# Poor Working Relationship between Doctors and Hospital Managers - A Systematic Review

**DOI:** 10.1101/2022.04.11.22273494

**Authors:** Collins Ogbeivor, Precious E Ogbeivor

## Abstract

**Background:** The problem of poor relationships between doctors and hospital managers is a common feature of many healthcare systems worldwide, including the United Kingdom’s NHS. Despite the significant impact that a poor working relationship between doctors and managers could have on the quality of care, there is limited research in this area.

**Objectives:** To investigate the organisational factors, contributing to the poor working relationship between doctors and hospital managers with a view to recommend potential solutions to address them.

**Methods:** We performed a systematic literature review; a comprehensive search of AMED, MEDLINE, CINAHL, plus with Full Text, SportDiscus and EBSCO EBooks from January 2000 to July 2019 and updated in March 2022, and no further article was found that meets the selection criteria. Mixed methods, qualitative studies and quantitative studies published in English language in peer reviewed journals between January 2000 and March 2022 were included. Study selection, data extraction and appraisal of study were undertaken by the authors. Quality criteria were selected from CASP Checklist.

**Results:** A total of 49,340 citations were retrieved and screened for eligibility, 41 articles were assessed as full text and 15 met the inclusion criteria. These include 2 mixed method studies, 8 qualitative studies, and 5 quantitative studies. A thematic analysis was undertaken, and narrative summaries used to synthesise the findings.

**Conclusion:** The findings of this systematic review show strong evidence of poor collaboration and lack of effective communication that contribute to poor working relationships between physicians and hospital administrators. The results from this review may guide the development of a hospital plan that involves both doctors and managers in the decision making process regarding the quality of patient care, which could potentially enhance the relationship between the two groups as it would build trust between them.

What is already known on this topic

- The problem of poor relationships between doctors and hospital managers is a common feature of many healthcare systems worldwide.
- Despite the significant impact this poor relationship could have on the quality of care and patient satisfaction, there is limited research in this area.

What This Study Adds

- We conducted a systematic review on the effect of tension between doctors and hospital managers on the quality of care provided in hospital or healthcare centres; 15 (qualitative, quantitative and mixed) primary papers were reviewed. This qualitative systematic study found considerable evidence of organisational factors that contributes to poor working relationships between doctors and managers.

## 1. Introduction

The problem of poor relationships between doctors and hospital managers is a common feature of many healthcare systems worldwide, including the United Kingdom’s (UK) National Health Service (NHS).^1, 2^ According to Powell and Davies^3^ good working relationships between doctors and managers are essential ingredients for the effective performance, safety and quality of the NHS. Therefore, a poor working relationship could have a significant impact on the quality of healthcare, as it could lead to high mortality rates, near misses, low staff performance as well as patient satisfaction.^2, 4-5^

Previous healthcare models involved government appointing hospital boards, made up of administrators, with members not necessarily part of the hospital community; e.g. former military officers or politicians with experience as public servants.^6^ However, one of the criticisms of this practice is that it was ineffective because it lacked competent technocrats who have the requisite knowledge and experience of long-term planning and proper management of hospital systems.^6^ With the growth of healthcare management and the emergence of physicians in hospital administration, the acceptance of this model among healthcare professionals has been further reduced.^6-7^ Spurgeon,^7^ also states that the involvement of managers, empowered to enforce government policy and the seeming conflicting role of clinical professionals e.g. doctors in hospital administration has led to tensions between the two groups This is corroborated by a study on doctor-manager relationships in both the US and the UK, which found that both groups agreed that relations between doctors and managers were poor;^8^ despite the obvious differences between the US and the UK system of healthcare delivery.

Despite the significant impact this poor relationship could have on the quality of care there is limited research in this area.^2, 4-5^ Also with the introduction of marketing into healthcare, i.e. the drive for increased efficiency, there is a well-established shift in public sector management for improved quality of healthcare, better clinical outcomes and improved patient satisfaction.^9^. The problem does not only persist, but it is likely to deteriorate in the coming years with the growing risks of doctors disengaging from management. To address this issue, we conducted a systematic review of literature on the evidence of poor working relationship between hospital managers and doctors with a view to identify the organisational factors that contribute to poor working relationship between doctors and hospital managers and suggest ways to overcome them.

## 2. METHODS

The qualitative systematic review defined by Ring et al.^10^ and the York Centre for Reviews and Dissemination^11^ guided the methodological protocol for this study. The review was carried out by consulting the following electronic databases: Ebscohost, AMED, MEDLINE, CINAHL, SportDiscus and EBSCO Ebooks from January 2000 to July 2019 and updated in March 2022, but no further article was found that meets the selection criteria. Reference lists from the relevant primary and review studies and grey literature as well as relevant healthcare management textbooks were consulted for information on manager-doctor relations.

The search strategy began with the use of multiple terms and key words that describe the population such as doctors, managers and physicians. These terms were linked together using the Boolean operator “OR” to ensure that articles retrieved contained at least one of the search terms. The same process was repeated for a second and a third set of terms related to the exposure (working relationships in hospital or healthcare service) and the study design (Mixed methods, qualitative studies and quantitative studied) respectively. These three sets of terms were then combined together with the Boolean operator “AND”. This allows for the retrieval of studies that are relevant to the study design and address both the population of interest and the exposure to be investigated. See Table 1 for detailed description.

**Table 1:** Quantitative Search - Combined Results of the CSP Electronic Database Searches of AMED, CINAHL, CINAHL Plus with Full Text, CSP Online Library Catalogue, eBook Collection (EBSCOhost), MEDLINE, SPORTDiscus.

| # | Search Terms | Combined Results from above Database Searches |
| --- | --- | --- |
| S1 | Doctors | 499,000 |
| S2 | Physicians | 1,546,371 |
| S3 | Physicians or doctors or clinicians | 2,214,757 |
| S4 | Medical doctors or practitioners | 1,406,777 |
| S5 | S1 OR S2 OR S3 OR S4 | 2,634,181 |
| S6 | Manager or managers | 318,119 |
| S7 | Manager or leadership | 561,491 |
| S8 | Manager or leader or executive or administrator | 881,949 |
| S9 | Hospital manager or managers | 318,119 |
| S10 | Hospital management or administration | 4,526,489 |
| S11 | Hospital directors | 2,162 |
| S12 | Trust management | 319 |
| S13 | Trust administrators | 18 |
| S14 | Trust managers | 321 |
| S15 | S6 OR S7 OR S8 OR S9 OR S10 OR S11 OR S12 OR S13 OR S14 | 5,246,569 |
| S16 | Poor relations or relationships | 2,776,115 |
| S17 | Conflict | 300,258 |
| S18 | Differences in opinion | 3,600 |
| S19 | Dispute | 120,739 |
| S20 | Disagreement or argument or conflict | 492,086 |
| S21 | S16 OR S17 OR S18 OR S19 OR S20 | 3,134,782 |
| S22 | Mixed method | 55,689 |
| S23 | Qualitative method | 38,649 |
| S24 | Quantitative method | 24,596 |
| S25 | Mixed or qualitative or quantitative | 1,821,590 |
| S26 | S22 OR S23 OR S24 OR S25 | 1,821,590 |
| S27 | S5 AND S15 AND S21 AND S26 | 49,340 |

### 2.1 Inclusion Criteria

Mixed methods, qualitative studies and quantitative studies that explored doctors and managers working relationships in hospital or healthcare service were included in this review. The settings of the included studies were hospital or healthcare services. Studies that were published in English language in peer reviewed journals between January 2000 and March 2022 were included. See Table 2 for details.

**Table 2:** Inclusion and Exclusion Criteria.

|  | Inclusion Criteria | Exclusion Criteria |
| --- | --- | --- |
| Population | Doctors and managers | Not doctors and managers |
| Exposure | Doctors and managers working in hospital or healthcare service | Doctors and managers not working in hospital or healthcare settings |
| Outcome | Studies on doctors and managers working relationships in hospital or healthcare service | Studies not centred on doctors and managers working relationships in hospital or healthcare service |
| Type of studies | • Mixed methods, studies, qualitative studies that are published appropriately<br>• Full texts of Studies<br>• Research studies in English Language or translation to English from other languages<br>• Studies with clear Ethical Approval | • Abstracts or summaries<br>• Commentaries<br>• Studies not in English Language<br>• Studies without ethical approval will not be included |

### 2.2 Exclusion Criteria

Studies were excluded if the target populations were not doctors (physicians) and managers (hospital administrators, executives, directors), who work in hospital or healthcare settings. Studies that were not focussed on doctors-manager relationships were excluded from this review. Studies that were not published in English language and before January 2000 were also excluded. See Table 2 for details.

### 2.3 Search Strategy and Search Outcome

A total of 49,340 citations were initially identified and retrieved from the Ebscohost electronic databases and additional 15 papers were also found from the reference lists and grey literature. There were 29,126 citations after removal of 20,229 duplicates. Of these, 29,085 articles were excluded based on the exclusion criteria and the remaining 41 articles were screened for their abstracts. Reference lists and grey literature were also searched, but no additional papers were found. Upon full text review of the 41 potentially eligible articles, 21 studies were excluded for the following reason; they were exploratory studies that described the relationships between doctors and nurses. 20 full text articles that were possibly relevant to this study were identified and reviewed for quality appraisal and five articles that were commentaries were excluded. (See Figure 1 below for details). 15 studies were included as part of the quality appraisal and synthesis.

**Figure 1:**
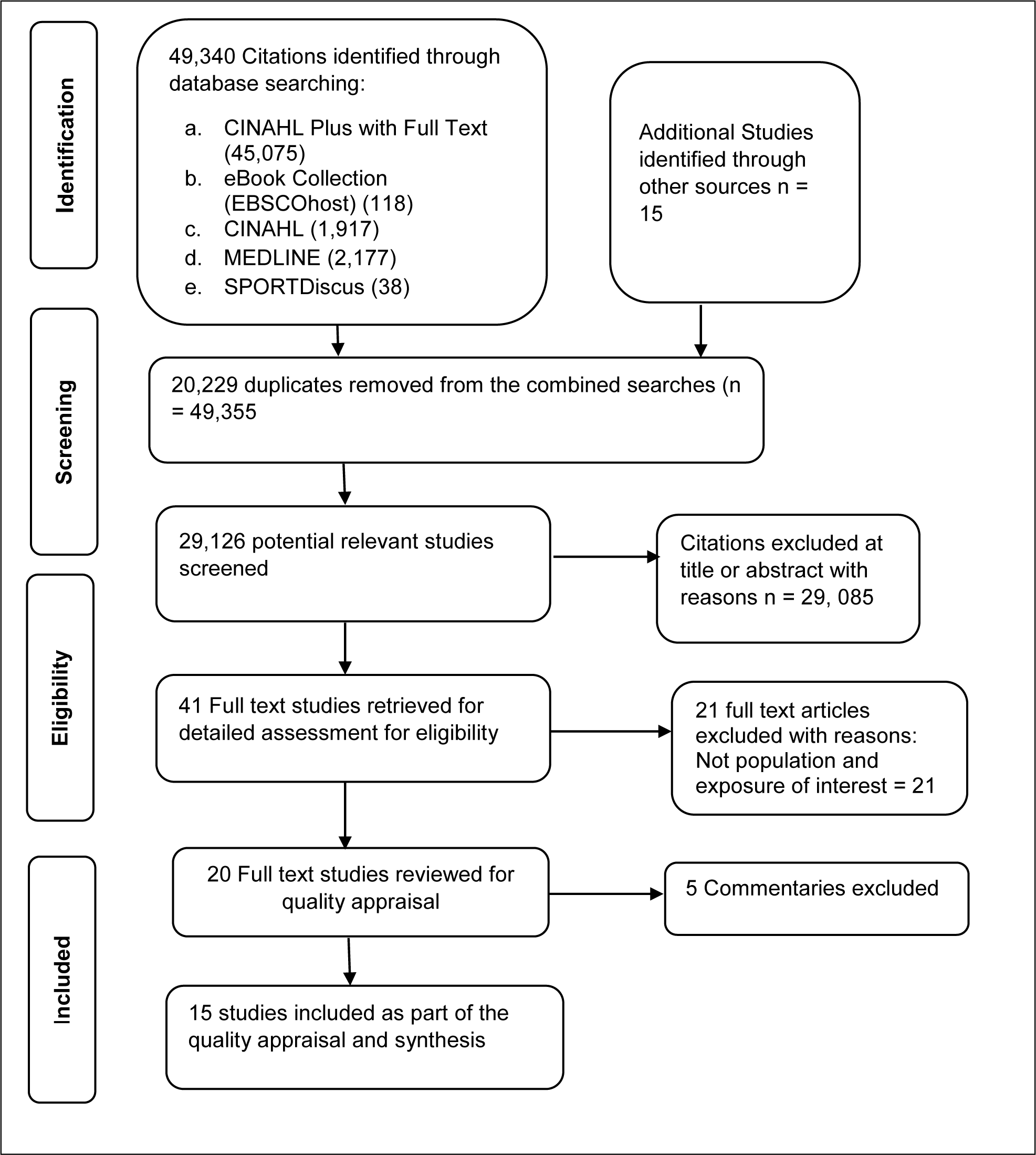
Proposed PRISMA Flow Diagram.

### 2.4 Quality Appraisal

Although it has been argued that quality assessment is not a major requirement for qualitative systematic review, however, it is recommended that studies that are retrieved should not have methodological issues.^12^ The quality appraisal of the studies that were included in this review were conducted using the Critical Appraisal Skills Programme^13^ Qualitative Research Checklist, a tool that has been developed and commonly used by researchers for checking the trustworthiness and rigor of qualitative research. The tool enables the assessment of a qualitative study’s aim, methodology, sampling process, data collection and analysis, ethics and findings. The tool contains 10 questions and each question was categorised as either ‘yes’, ‘can’t tell’ or ‘no’. If one question was scored ‘yes’, it was counted as 1 point. If all questions were assessed as ‘yes’, its total score is 10. The total quality score for a study is a maximum of 10 points. If the question was assessed, as ‘can’t tell’ or ‘no’ it was counted as 0. The researchers conducted the quality appraisal to ensure that all the studies included in the review had adequate methodological rigor. After the quality assessment, all the 15 studies selected for full review had a quality score of 8 points or more.

### 2.5 Data Extraction and Synthesis

A data extraction form by Bethany-Saltikov^14^ was used as a data registry and as a guide for identification of poor working relationships between doctors and managers. Details of the author, year of publication, purpose of the study, study design, setting, population, exposure and outcomes were included in the data extraction form. This qualitative systematic review adopted the Ring et al.^10^ thematic synthesis of qualitative findings. It involves identifying and coding recurring concepts from the selected studies’ textual findings, synthesising the codes into themes, and generating higher level themes. This enable the authors to gain an overview and make sense of the data, and also manage, synthesise and interpret the data in a structured and systematic manner using descriptive and illustrative accounts. See table 3 for details of codes and synthesised themes.

**Table 3:**
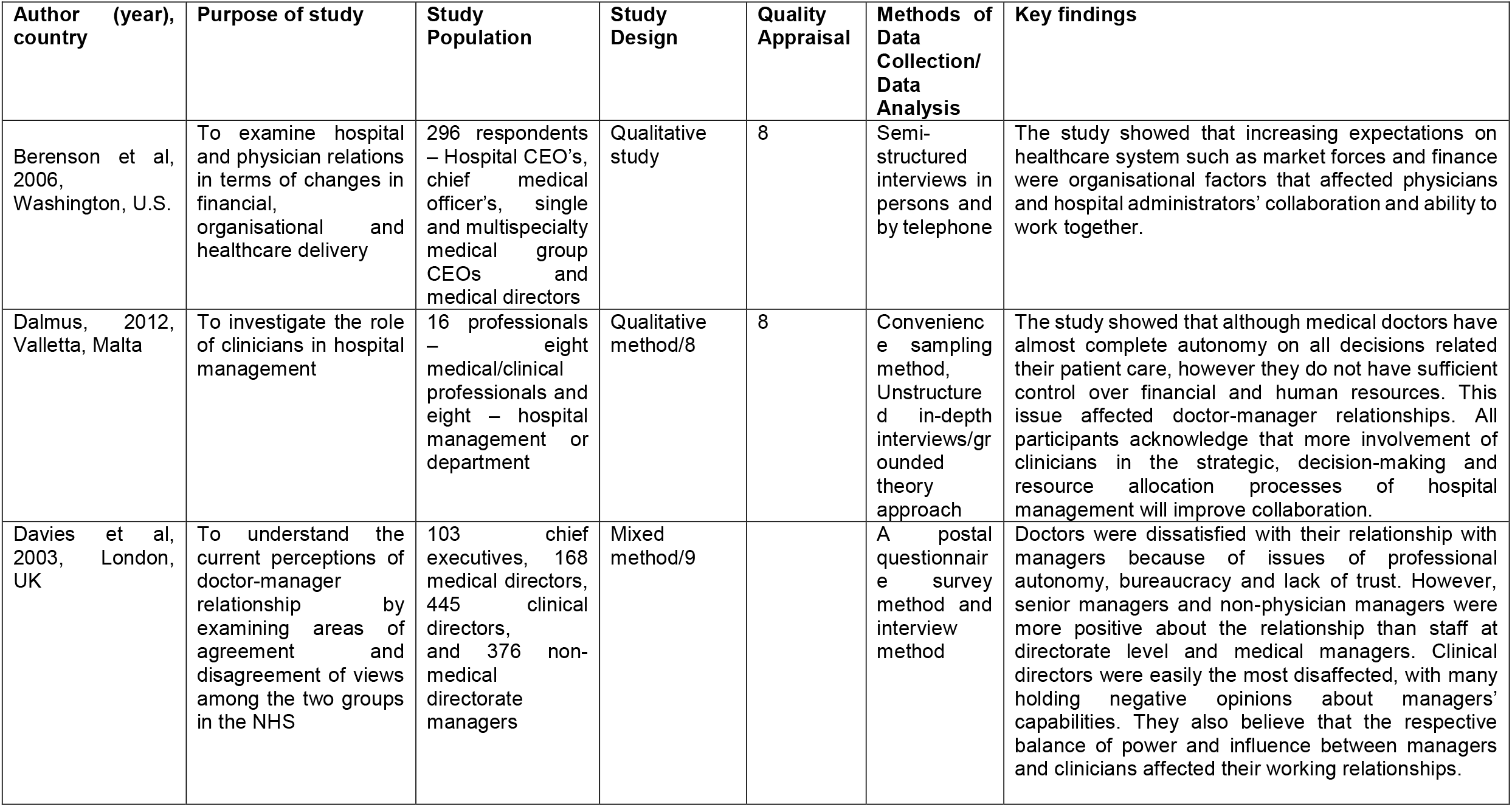
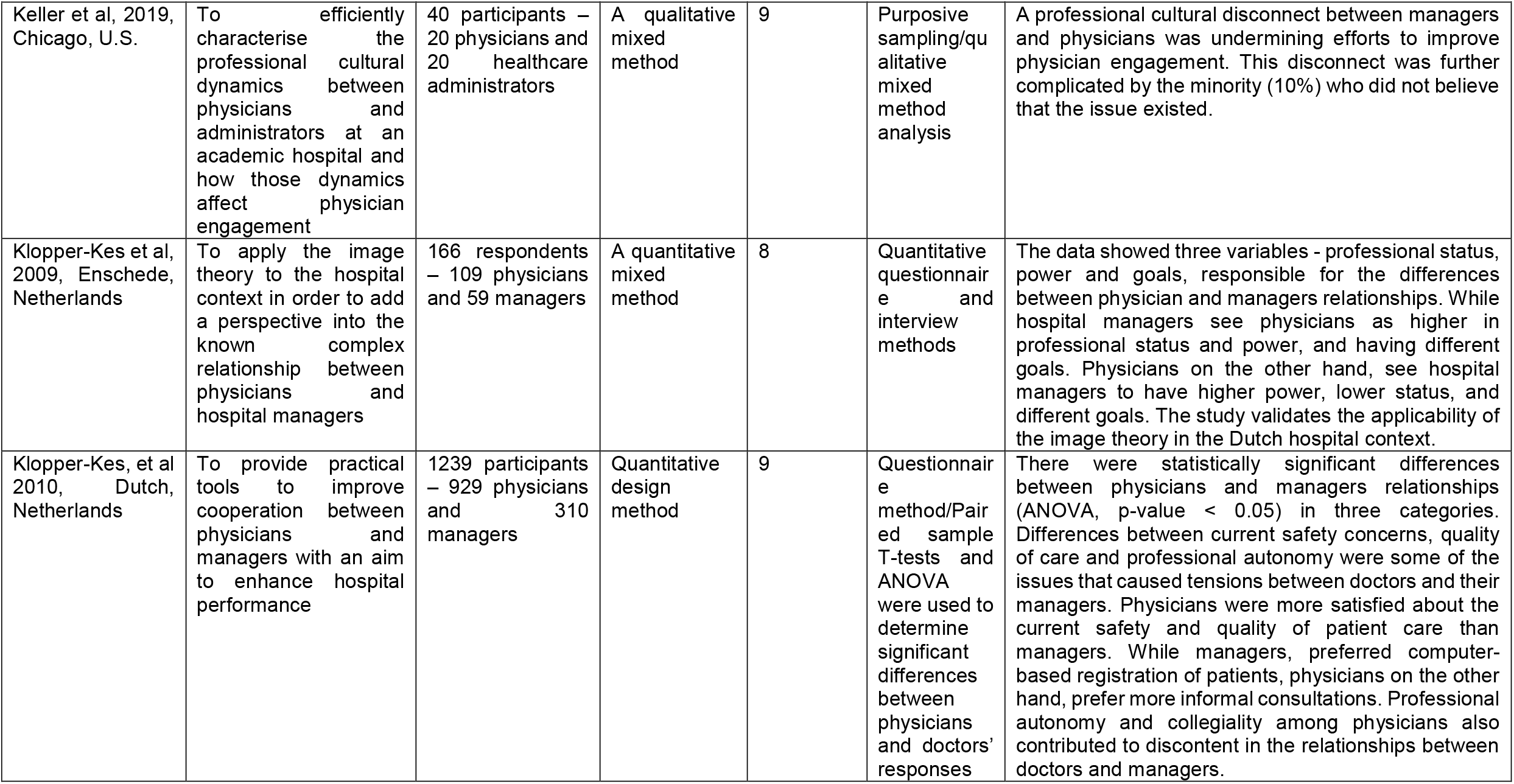
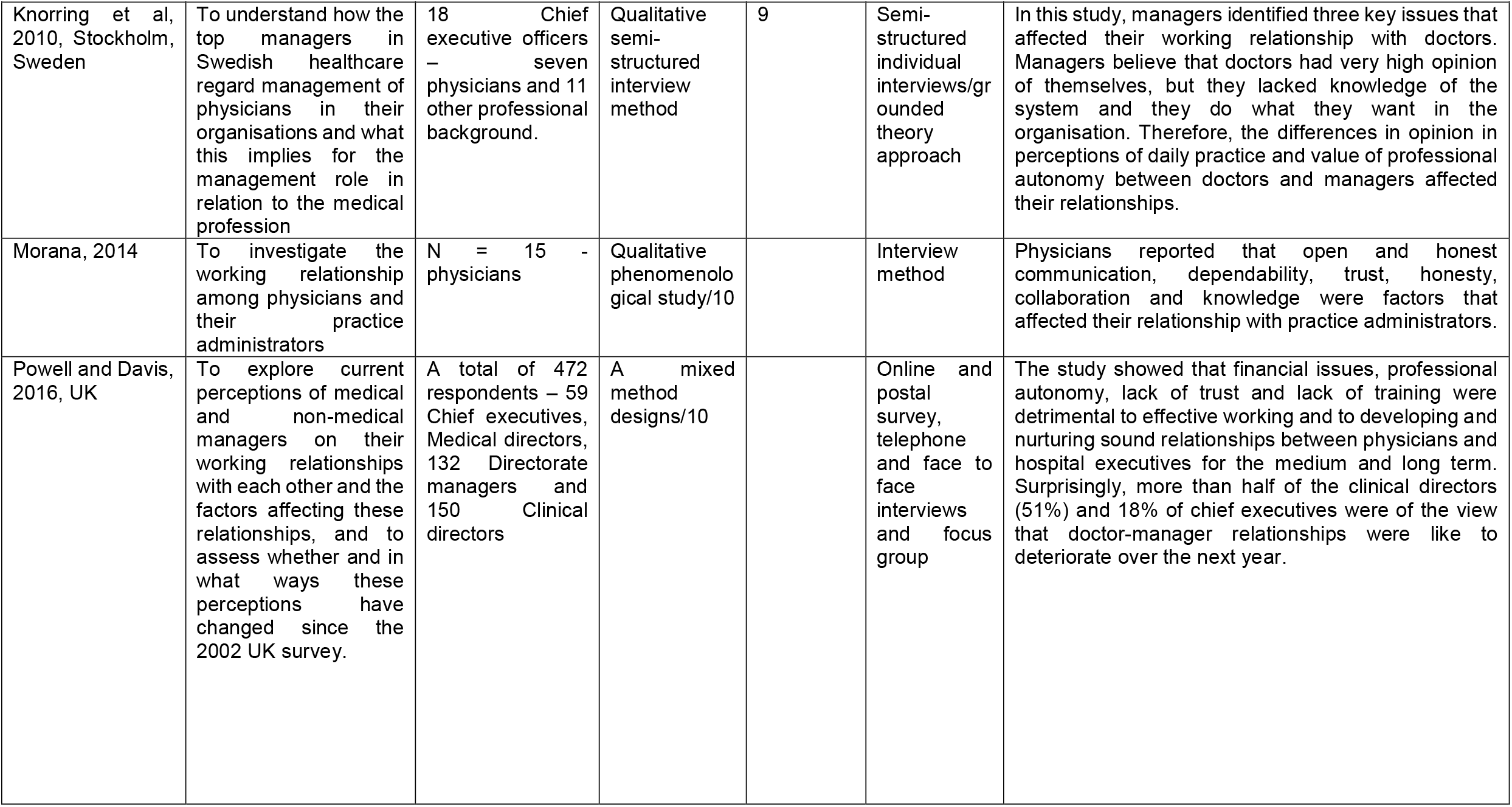
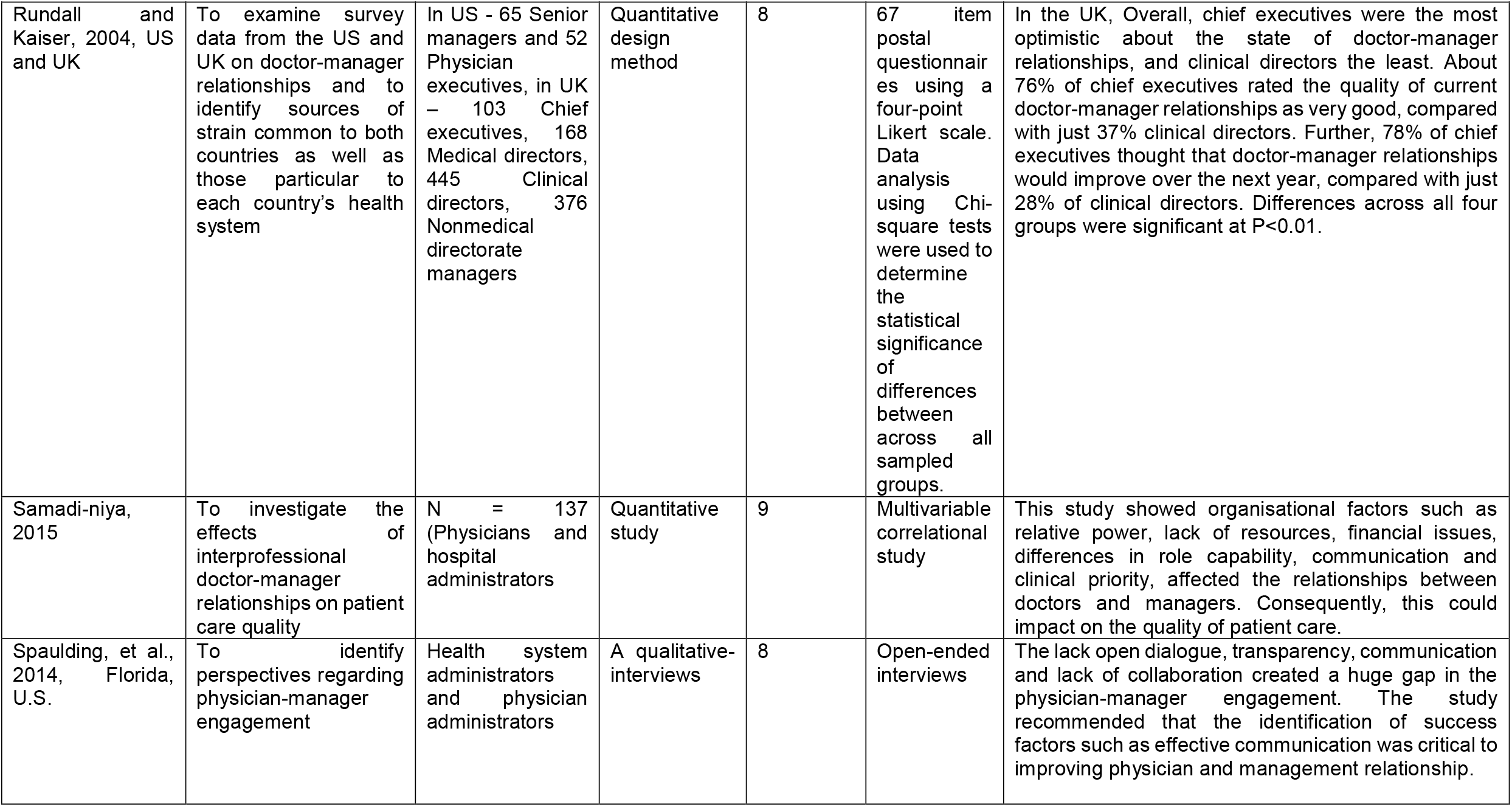
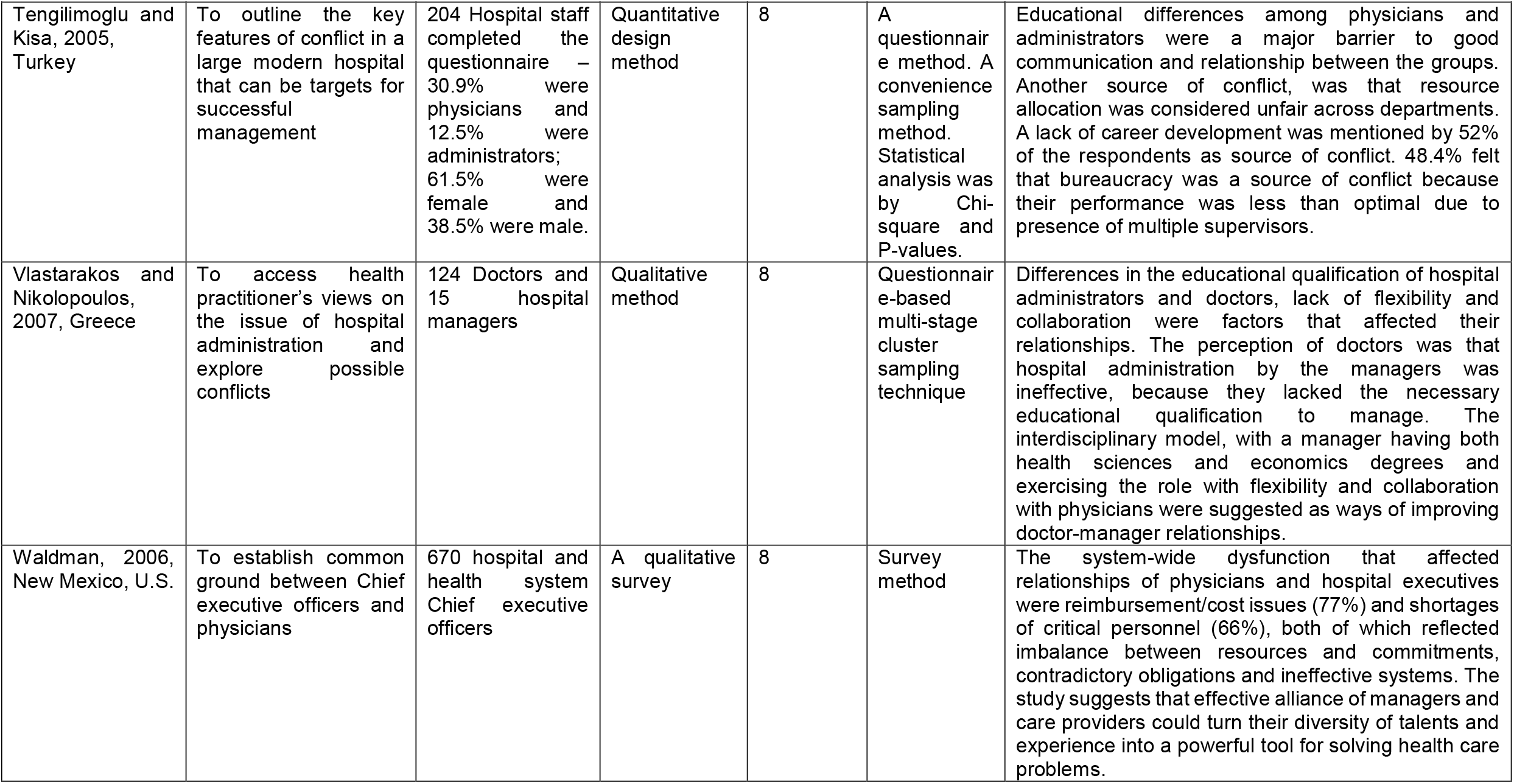
Summary of the Included Studies.

## 3. RESULTS

Fifteen peer-reviewed journal articles were included in this systematic review. Six studies discussed factors affecting doctor-manager working relationships.^8, 15-19^ Four studies explored perceptions of physicians-managers relationships and discussed their different viewpoints.^3.20-22^ One study focussed on the involvement of physicians with hospital administrators in hospital management.^23^ Two studies focussed on work-related conflicts between physicians and managers relationships.^6,24^ One study investigated the role of educational qualifications between medically educated and managerially educated senior manager relationships.^25^ One study explored the cultural dynamics between physicians and hospital administrators.^26^ Two studies were conducted in the UK, five in the US, one study was conducted in both the UK and the US, two studies were from the Netherlands, one study each in Malta, Sweden, Norway, Turkey and Greece. Four studies were quantitative, seven were qualitative and four used mixed methods.

See Table 3 below, which summarises all the studies included in this review. The studies’ details, design, samples, data collection, data analysis and key findings were summarised in the table. Five key themes were identified from the data analysis (see Appendix 1 for details of the process for data extraction using thematic approach) and these are related to organisational factors that caused poor doctor-manager relationships (see Table 4). These key themes and sub-themes are discussed in the next session below.

**Table 4:** Summary of Thematic Analysis: Organisational Factors Causing Poor Doctor-Manager Relationships.

| Main Themes | Code in the texts |
| --- | --- |
| Poor collaboration between managers and doctors | “We need to have open dialogue, transparency, communication, not develop a ‘we and them’ type relationship” (Powell and Davis, 2016, Spaulding, et al., 2014)<br>Competition as a potential source of disagreement between managers and doctors (Berenson et al, 2006)<br>There needs to be more partnering and more physician driven models (Spaulding, et al., 2014)<br>Without involving the physicians in defining that positive environment, the organisation runs the risk of developing wrong model (Spaulding, et al., 2014)<br>Management structures, which focus on the patient rather than on professional hierarchies (Dalmas, 2012)<br>Disconnection between the board and divisional or doctorate level (Powell and Davis, 2016)<br>Lack of development initiatives for cross-professional collaboration (Dalmas, 2012)<br>Communication issues (Davis, et al., 2003, Morana, 2014, Spaulding, et al., 2014)<br>Engagement survey (Keller, et al., 2019)<br>Trust, respect and shared values and objectives (Dalmas, 2012, Morana, 2014)<br>Bureaucracy- presence of multiple supervisors (Tengilimoglu and Kisa, 2005) |
| Cultural issues | Culture of medicine versus culture of management (Samadi-niya, 2015, Keller, et al., 2019)<br>Cultural views of managers are business and profit oriented, while doctors’ views are clinical and patient focussed (Morana, 2014, Samadi-niya, 2015)<br>Both managers and doctors showed differences in perceptions of daily practice (Klopper-Kes, et al, 2010)<br>Differences in physicians’ and administrators’ professional backgrounds, values and thought processes (Keller, et al., 2019)<br>Administrators’ and doctors’ differences in loyalty to organisation and profession (Keller, et al., 2019) |
| Power and autonomy | Physicians think hospital manager are pushing the limits by trying to go as far as possible (Klopper-Kes, et al., 2009)<br>The influence of the trust board (Powell and Davis, 2016)<br>Physicians see hospital managers as threat to their status and power, and vise versa (Klopper-Kes, 2009)<br>Hospital managers think physicians ruthless and try to stay in power as long as they are the biggest and strongest (Klopper-Kes, et al., 2009)<br>Lack of proper and clear definition of roles and responsibilities (Dalmas, 2012)<br>Doctor-manager differences in value of professional autonomy (Davis, et al., 2003, Klopper-Kes, et al, 2010)<br>Disagreement on the relative power and influence between management and physicians (Rundall and Kaiser, 2004, Samadi-niya, 2015) |
|  | Management exert pressure on physicians to discharge or transfer patients early (Rundall and Kaiser, 2004)<br>CEO's thought physicians were reluctant to abide by rules, avoid participating in group meetings (Von Knorring, et al., 2010)<br>"Half of administrators and physicians oriented themselves as *bosses* and *islands*" (Keller, et al., 2019)<br>Non-medical managers were perceived to hold all of the power (Powell and Davis, 2016) |
| Finance and resource issues | Competition over services between doctors and managers (Berenson et al, 2006)<br>Increased public expectation for improved patient safety and quality of care (Berenson et al, 2006, Dalmus, 2012)<br>Physicians are asked to do more for less pay (Samadi-niya, 2015)<br>The use of hospitalists rather than physicians and specialists (Berenson et al, 2006)<br>Management is driven more by financial than clinical priorities (Powell and Davis, 2016, Rundall and Kaiser, 2004, Tengilimoglu and Kisa, 2005, Samadi-niya, 2015)<br>Financial arrangement of hospitals and physicians with payers (contract) (Samadi-niya, 2015)<br>Adequacy of resources (Waldman, 2006 and Samadi-niya, 2015) |
| Educational differences/challenges | Differences in educational qualification of doctors and managers (Tengilimoglu and Kisa, 2005, Vlastarakos and Nikolopoulos, 2007)<br>Impact of training on relationships between senior clinicians and management (Powell and Davis, 2016)<br>Educational differences led to communication problems between different professionals (Tengilimoglu and Kisa, 2005)<br>Lack of development initiatives for cross-professional collaboration (Dalmus, 2012)<br>Training in management skills (Dalmus, 2012)<br>Lack of opportunity for career development (Tengilimoglu and Kisa, 2005)<br>Physicians lack knowledge of the system (Von Knorring, et al., 2010)<br>Physicians do not respect opinion of managers with education in history or geography (Samadi-niya, 2015) |

## Organisational Causes of Poor Doctor-Manager Working Relationships

### Theme 1: Poor collaboration and communication

Nine of the studies reviewed, reported lack of collaboration and communication as organisational factors affecting the relationships between physicians and hospital managers.^3,15,18-20,22-24,26^ Furthermore, three studies^3,18,22^ found that lack of open dialogue, transparency, communication resulted in a ‘we versus them’ type of relationship between doctors and hospital administrators. In the study by Spaulding, et al.^22^, one of the hospital administrators had this to say: “I think we need to do a better job of listening to our physicians…not just listening to them, but really hearing them…what their core values are, and engaging with them.”^22^ Equally, Samadi-niya^19^ found that lack of teamwork and communication has significant impact on inter-professional relationships between the two groups.

Bureaucratic involvement of multiple supervisors^24^ lack of developmental initiatives for cross-professional collaboration, trust, respect and shared values and objectives were identified as some of the barriers to physician-administrator rapport.^18,23^ This point was re-echoed by Weiner, et al.^27^ stating that lack of collaboration does not only have a negative effect on inter-professional relations between the two groups, it also hinders the improvement in the quality of patient care.

### Theme 2: Cultural Issues

Three other studies described cultural issues as barriers to relationships between doctors and managers.^16,18-19,26^ Keller, et. al.^26^ reported that physicians’ and administrators’ professional backgrounds, values and beliefs differed considerably. Furthermore, the researchers reported that the differences in physicians’ and administrators’ professional backgrounds, values and beliefs affected their working relationships. For example, while administrators believe that excellent patient care can be achieved by promoting the organisation and its brand, physicians on the other hand are of the view that excellence in patient care was attainable by advancing profession/specialty through education and research.^26^

Another key cultural difference that affected the relationships between the two groups was their different approaches to decision making. The physicians’ viewpoint was that patient care occurred in high-acuity, with short clinical decision-making time, and where a lot of information is shared in a single best course of action.^26^ On the contrary, administrators considered organisational care with a relatively much longer time and information dissemination involving multiple channels.^26^ These views compared favourably with Bujak,^28^ who reported that “physicians have an expert culture and administrators have an affiliative culture”. According to Samadi-niya,^19^ the cultural views of managers are business oriented, rooted on profitability, while physicians have dissimilar cultural views, which are clinical and patient focused.

### Theme 3: Power and Autonomy

Nine studies cited the complexity of power and autonomy as a barrier to doctor-manager relationships.^3,8,16,20-21,23,26,29^ Physicians saw hospital administrators as being higher in power and hospital administrators see doctors as being higher in power^21^ This implies both groups feel relatively “powerless” in the same organisation and the practical implication of this is that there could be lack of proper and clear definition of roles and responsibility in achieving organisational goals such as improved quality of patient care and staff performance^21-22,^ In one of the studies, a hospital administrator was noted saying “if they should know what I can offer them, and know what kind of things they could use me for, our relationship and cooperation would not be such a problem”.^22^

Doctor-manager differences in value of professional autonomy was another reason cited as a barrier to a harmonious working relationship between the two groups.^17,21-22^ For example, hospital administrators described how doctors were reluctant to abide by rules, avoided participating in group meetings with them, and in many respects, chose to follow their own agendas.^17^ This type of “do-what-you-want” mentality was perceived by the administrators as “strong” and not limited to clinical matters. Similarly, Keller, et al.^26^ reported that “half of the administrators and physicians interviewed described their relationship as “*bosses”* and “*islands”* where increasing communication between them meant “getting them on-board” or “making them understand” and presence was about policing the activities of others*”*.

### Theme 4: Finance and Resource

In seven studies, financial and resource challenges were reported as barriers to relationships between doctors and managers.^3,8,15,19,23-25^ A directorate manager in the study done by Powell and Davis^3^, cited the negative impact of financial and resource constraints on relations between the two groups, stating that “*the increasing financial constraints and increasing demands on the service are taking their toll on all relationships*” (p.25). It was noted that both the physicians and hospital administrators agreed that the bond between them is negatively affected by the nature of financial targets set by the funding providers.

Four studies^3,8,19,24^ found that part of the conflict and disengagement between the two groups was because doctors felt management was driven more by financial gain rather than clinical priorities. Increased public expectation for improved patient safety and quality of care in the face of financial scarcity was identified as another source of tension between physicians and managers.^15, 23^

### Theme 5: Educational Differences/Challenges

Four studies cited differences in educational qualifications of doctors and managers as a source of tension and lack of engagement between the two groups.^6,15,23-24^ For example, lack of management training for doctors and executive coaching on leadership style could hamper the relationship between doctors and managers.^3^ Hence, joint training events for the groups have been shown to improve their collaboration.^3^ In the study by Vlastarakos and Nikolopoulos,^6^ 61% of the doctors working in the hospitals ignored the basic degree of the hospital manager, while 71% of the doctors felt the degrees were inadequate for the efficient management of the hospital. Furthermore, Tengilimoglu and Kisa,^24^ concluded that educational differences between physicians and administrators were a major barrier to effective collaboration and integration between the groups. Similarly, it has been stated that through professional training, regulation, medical licensing and certification, physicians have this communal type relationship within the hospital, which Kaissi,^29^ termed “occupational community”. This occupational community relationship among doctors influence their interaction with hospital managers who on the other hand are not viewed as part of that community because they are individuals from various educational backgrounds such as business, public administration and accounting.^29^

## 4. DISCUSSION

This qualitative systematic study found considerable evidence of organisational factors that contribute to poor working relationships between doctors and managers. This review identified five major themes from the studies that were reviewed. The first was poor communication and collaboration amongst physicians and hospital administrators. Several authors have reported that there are well known challenges in the communication and group work between hospital executives and doctors.^2,29-31^ In this review, respondents highlighted lack of open dialogue, transparency, communication as factors that created a rift in the relationship between doctors and hospital administrators. Doctors felt that their inability to access hospital executives created a “we versus them” adversarial type relationship.^3,32^ Doctors also felt they were not being listened to by the hospital executives.^3^

Previous research in healthcare settings ^3,18,23,29,33-34,36^ suggests that if there is a specific plan, concentrated effort and resources in creating and maintaining effective working relationships between different groups such as doctors and managers working within healthcare services, communication and collaboration between them is likely to improve. The practical implication of such a strategic plan is that, not only will the different groups agree on key issues that affect service provision but there will also be enhanced cooperation and collaboration in achieving set objectives.^21^

Cultural issues were the second theme cited by majority of the studies included in this review. It has been reported that cooperation and communication between physicians and managers are affected by differences in their professional and organisational cultures.^16,29^ Furthermore, differences in organisational values, views and aspirations between physicians and hospital administrators were reported as obstacles for successful relationships between the groups. Although both doctors and managers agree on guaranteeing the safety of patients and improving their quality of care, they disagree on the level of involvement in the implementation.^16^ This disagreement is based on differences in meaning, values, and behavioural norms which are generally not comparable by the same standards.^29^ For instance, the physicians’ primary loyalty is to their patients, while managers have a strong allegiance to the organisation they serve.

The different socialisation and training that managers and physicians receive results in different worldviews, value orientation and expectations, which can hinder harmonious relationships between them.^16,29^ However, if these differences in perceptions are recognised and harnessed, they can become a veritable tool in enhancing their relationship, more so that survival in the current health care environment requires a diversity of skills, orientations and thought processes.^29^ This is consistent with the suggestion by Brockschmidt^37^ advising that organisations should adopt a corporate culture that allows both physicians and hospital managers to play important roles in solving conflicts of views, values and behavioural beliefs between them. One of the strengths of his suggestion is that the cultural divide between doctors and managers regarding business profitability and patient centred care could be a potential source for discussion and corporate engagement between the two groups.

The third theme identified was power and autonomy. In the studies under review, physicians viewed administrators as superiors with higher administrative powers, while managers perceived doctors as being higher with clinical decision-making powers. These perceived differences in professional autonomy and power does not only create tensions that can sometimes be counterproductive to the attainment of shared objectives but can also negatively affect the relationship between the two groups.^38^ According to Klopper-Kes, et al.^21^ if hospital administrators and physicians understand clearly each other’s roles and responsibilities in achieving organisational goals such as improved quality patient care and staff engagement, any perceived differences between the two groups could become key strengths in their relationship.

This review highlighted the fact that physicians, compared to hospital administrators were more focussed on clinical autonomy – that is taking independent decisions on patient care, whereas hospital administrators were more concerned about organisational bureaucracy and accountability. While physicians are patient-oriented, practicing their specialty well and treating more patients, they are easily frustrated by organisational bureaucracy.^2,38-40^ On the other hand, hospital managers are mindful of managing the organisation, balancing the needs of specialty areas and physicians against each other, in the face of declining revenues.^2,38^ These differences create tensions in their working relationships.

Another significant challenge to physicians’ autonomy is the increasing pressure from governments and hospital executives for them to be transparent and systematic in aspects of their clinical work such as scheduling, follow-up and communication.^2,20,38^ Therefore, Edwards^2^ recommended that both physicians and hospital administrators should develop guidelines, protocols, and develop the use of information to feedback utilisation data, cost effectiveness and clinical outcomes. In addition, it has been suggested that mutual respect for physician-hospital manager differences, responsible autonomy between the two groups, avoiding personal attacks and keeping to the principles of shared decision making – particularly in difficult areas such as resource control and accountability, could potentially improve relations between doctors and hospital administrators.^2,22,34-35^

The fourth theme identified in this qualitative systematic review was related to finance and resource challenges. Doctors and hospital managers/directors do not only face significant financial challenges, they also struggle to align behaviours to achieve cost and quality goals in today’s healthcare environment.^38^ Several authors have cited the role of administrators in the management of hospital resources as financial bookkeepers.^2,8,40^ However, this role may affect physician-administrator relationships as doctors do not accept the accounting mind-set of managers, as this may suggest critical evaluation of their practice.^6,8,40-41^ This implies that for hospital administrators to achieve efficiency in the services provided by doctors, they need to adopt a management style that is flexible, which takes into account the widest consent of all healthcare professionals such as medical doctors.^42-43^

The final theme identified by this review was educational differences/challenges between doctors and hospital executives/managers. This systematic review found that majority of doctors felt that the hospital administration is ineffective because the hospital managers do not have a health sciences degree.^6^ By way of resolving these issues some researchers have recommended a combination of medical doctor/master’s degrees in business administration training programmes or a post graduate training programme in healthcare administration for healthcare professionals such as physicians and hospital executives.^40,44^ This suggestion resonates well with the statement made by Kaissi,^29^ that more and more physicians are taking business courses and acquiring master’s in business administration (MBA) degrees in order to become a physician executive, however once they attain this role, their loyalties shift from their colleagues to that of the organisation. This shift in loyalty by the physician-administrator negatively affects their relationship with other practicing physicians.^29^ Conversely, Chhetri^32^ argues that because doctors share a common educational and professional background, they naturally respect and trust other physicians including those in administrative positions, compared with non-clinical hospital executives with different educational and professional experiences. These differences between practising doctors and non-physician managers create a great difficulty in reaching mutual understanding regarding the process of healthcare delivery and quality improvement.^32^ This suggests that hospital administrators need to pay enough attention to a mutual but different viable educational and career development path for both doctors and hospital managers.^44-46^

## 5. METHODOLOGICAL ISSUES

One of the limitations of this study is that there are few primary UK studies on poor working relationship between doctors and hospital managers, therefore this review looked at this issue from a global perspective. Also, by considering only English-language articles, we may have excluded other relevant studies. Despite these limitations, this review suggests several implications of poor working relationships between physicians and hospital administrators and has provided some solutions to resolve them in a manner that is sustainable.

## 6. IMPLICATION FOR PRACTICE & RESEARCH

To our knowledge, this systematic review is the first qualitative synthesis study to explore organisational barriers to cordial working relationship between doctors and managers. Based on the challenges identified in the studies under review, it was recommended that a hospital governance plan that involves both doctors and managers in the decision-making process regarding the quality of patient care, could potentially enhance the relationship between the two groups as it would build trust between them. It is also recommended that recognising and harnessing the differences such as diversity of skills, orientations and thought processes that exist between the two groups and using these as a viable tool in improving their relationship. The studies did not use any theoretical framework to conceptualise the psychosocial factors of intergroup relationships such as those involving doctors and hospital managers. It is assumed that a theoretical model that considers the social and psychological aspects of inter-communication between doctors and managers could have helped to understand the problems better. Therefore, future research should consider these aspects because solutions could be easier when the problems are investigated through a theoretical lens.

## CONCLUSION

In summary, this study found that better communication, an understanding of the different cultural issues affecting doctors and hospital managers, as well as greater involvement of both groups in decision making among other things will go a long way to ease the tensions in the working relationship between the two groups.

## Data Availability

None

## Acknowledgements

The authors wish to Dr Senaka Fernando for his profound supervision, guidance, support and encouragement for this research.

## Ethical approval

Not required.

## Contributors

PO conceived and directed the study. Both authors contributed to the design of the study. PO and CO managed data acquisition. CO provided expert advice on the analyses. CO drafted the manuscript. CO provided additional important intellectual and substantial advice throughout the study and on all redrafts of the report. PO is guarantor for the study.

## Appendix 1: Summary of Main and Subthemes of Included Studies

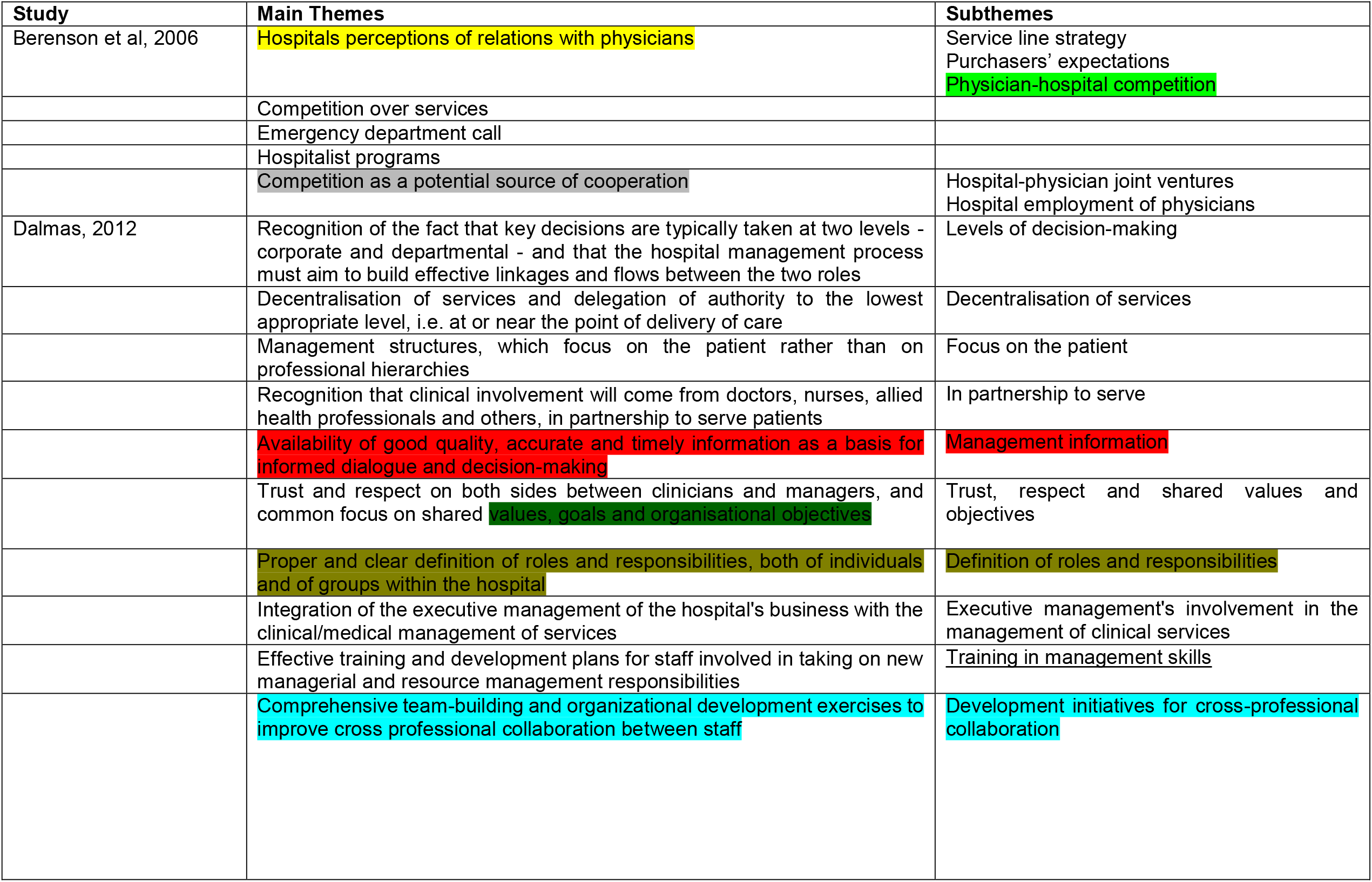
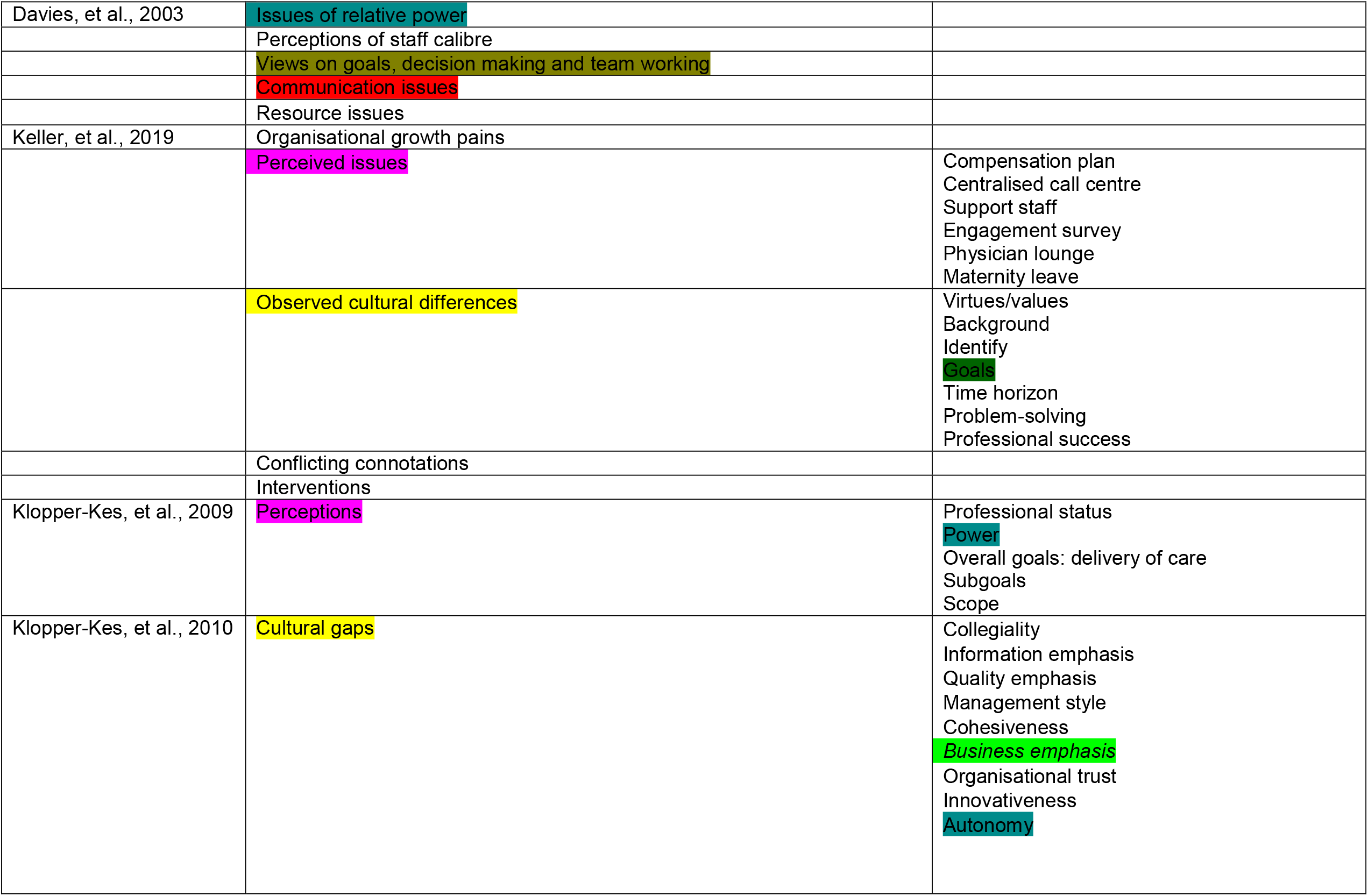
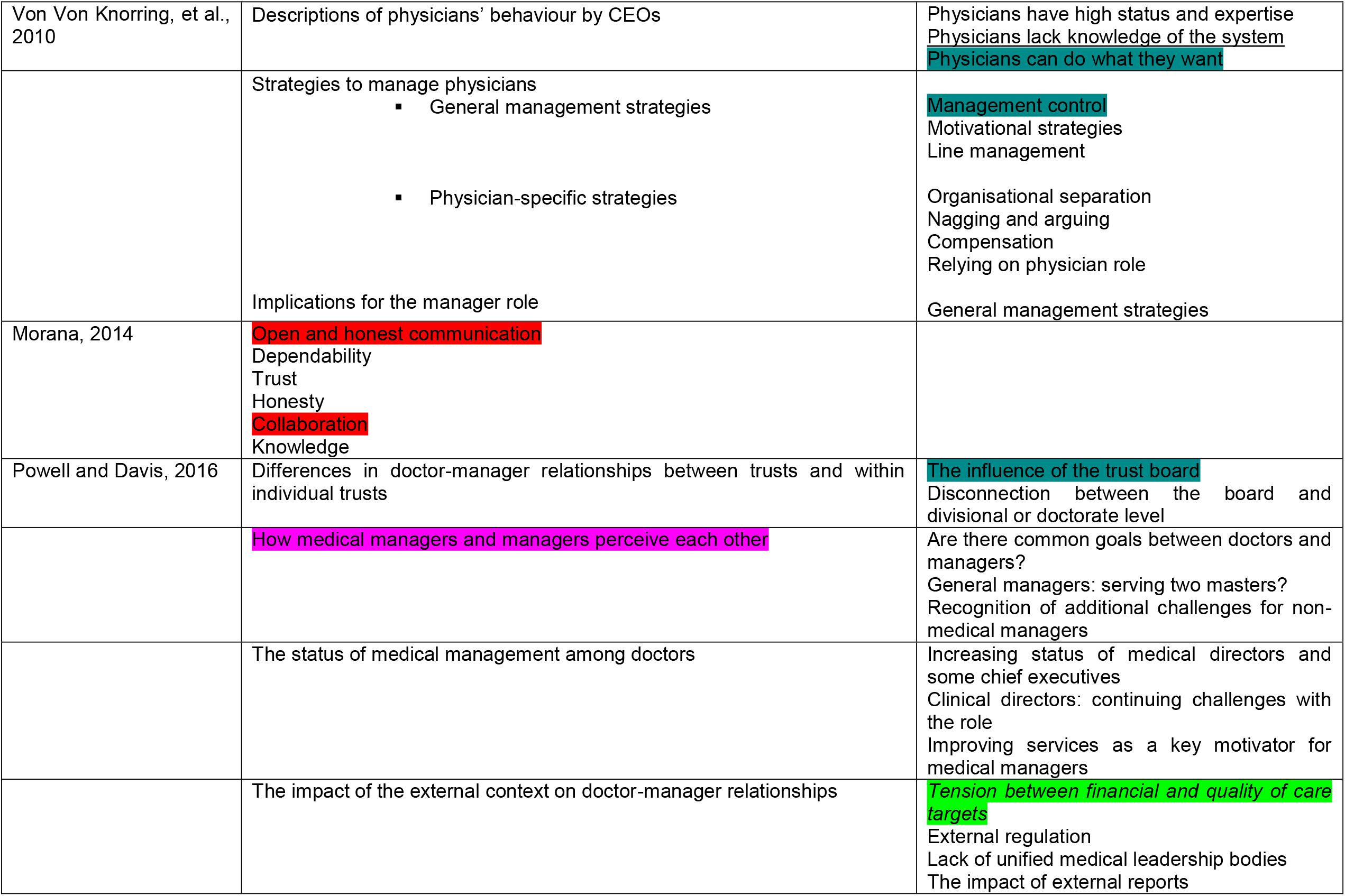
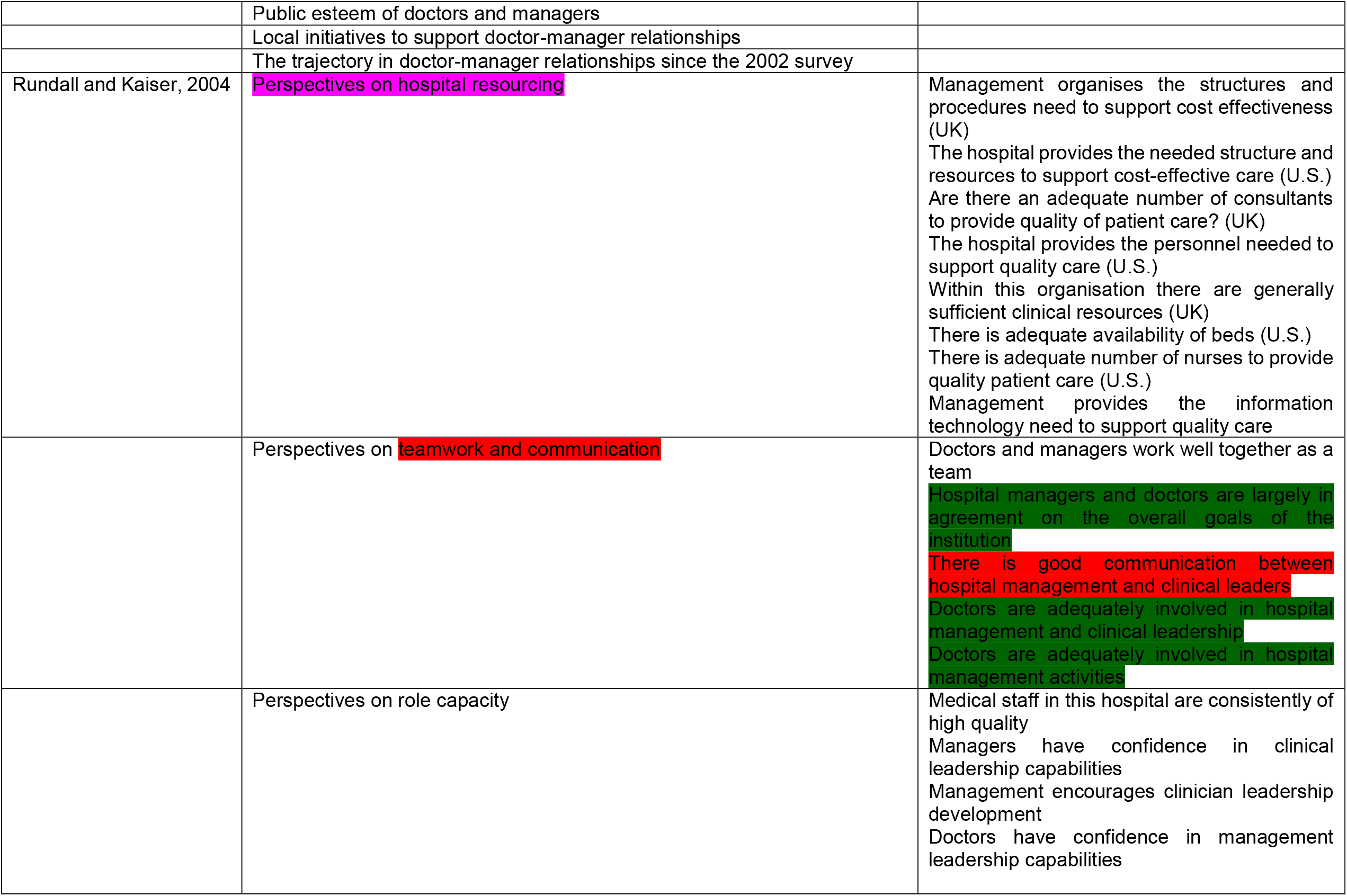
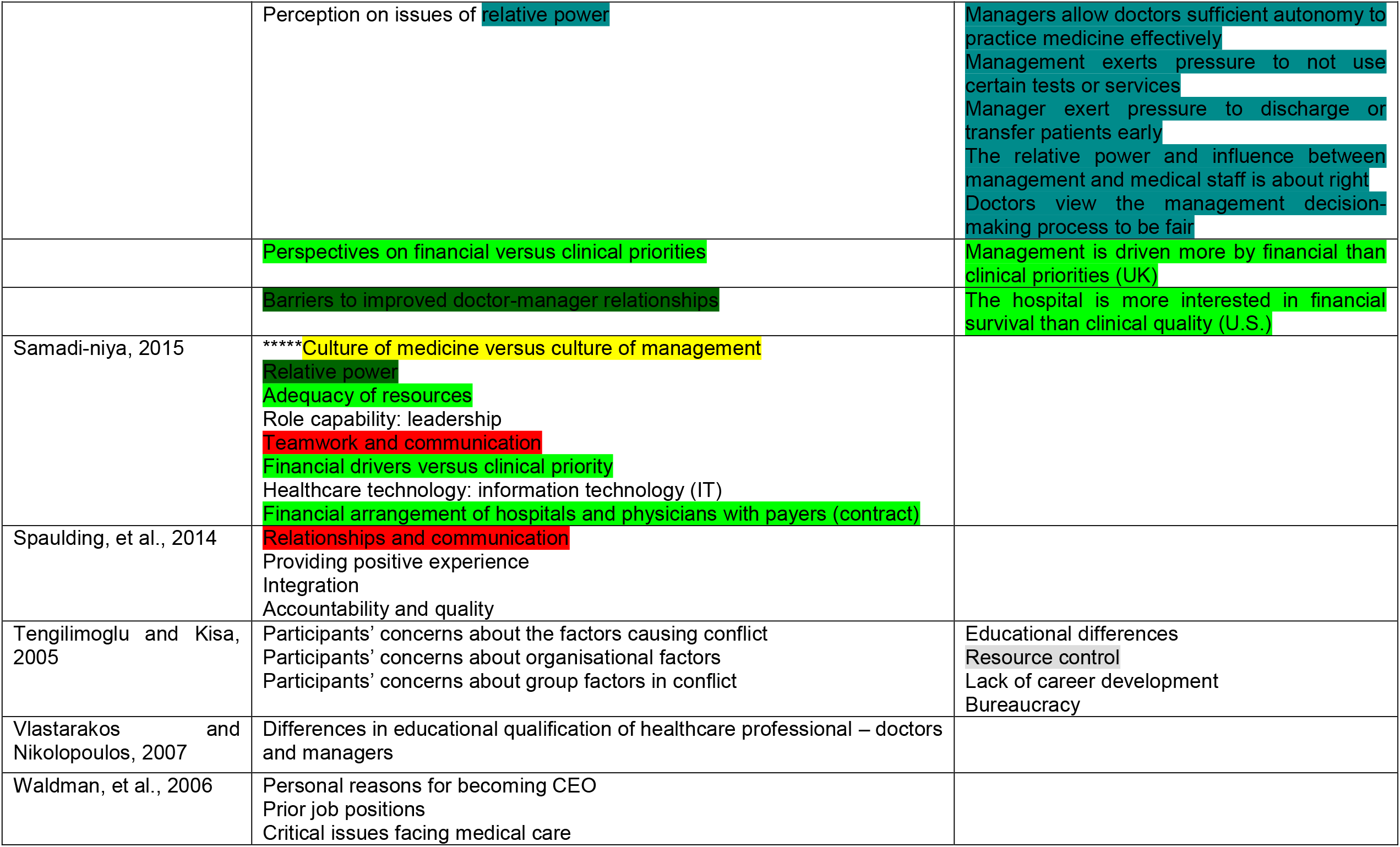

